# Proteo-transcriptomic analysis reveals brain-region-specific gene and biomarker expression in post-mortem neural tissue of Alzheimer’s patients

**DOI:** 10.1101/2022.11.26.22282736

**Authors:** Gokulanath Mahesh Kumar

## Abstract

Alzheimer’s disease is a neurodegenerative disorder, marked by dementia, impaired judgment, and a decline in cognitive abilities, that affects over 50 million people worldwide. The etiology of the disease is influenced by a myriad of genetic and environmental factors including APOE4 allele status and sex and can be characterized by a variety of neuropathological markers, such as amyloid-beta (Aβ) plaques and neurofibrillary tangles (NFTs) in specific areas of the brain, as well as the dysregulation of certain genes. However, how APOE4 status and sex affect the expression of these biomarkers and the gene dysregulation patterns specific to each brain region have yet to be determined. In this study, statistical and correlational analyses were performed to identify and quantify the relationship between the expression of proteo-transcriptomic biomarkers associated with neurodegenerative disorders and neuronal development in the frontal white matter, parietal cortex, temporal cortex, and hippocampus with patient demographic and genetic information. Further analysis determined whether the observed correlations were brain-region-specific or sex-specific. There was a statistically significant relationship between APOE4 status and degree of Aβ accumulation (P < 0.001) and neurofibrillary tangle formation (P = 0.04). Aβ plaque deposition was found to be more severe in females (P = 0.001) and the intensity of Aβ and NFT formation differed across brain regions (P < 0.001, P = 0.001 respectively). Furthermore, transcriptomic analysis revealed brain-region-specific gene dysregulation patterns. These patterns can identify therapeutic targets for future targeted gene and protein therapies in specific brain regions to treat Alzheimer’s disease.

## Introduction

Alzheimer’s disease is a neurodegenerative disorder marked by progressive detriment to cognitive and memory capabilities^1^. Age is the major risk factor, with symptoms of memory loss usually appearing in the mid-60s in those with late-onset of AD (LOAD). The dysregulation of various genes plays an essential role in the etiology and pathogenesis of Alzheimer’s disease. Most notably, dysregulation of amyloid precursor protein (APP) and its proteases: β-secretase, γ-secretase, and presenilins result in the miscleavage of APP and production of amyloid β peptides.

These prions aggregate to form extracellular amyloid β plaques, especially in the gray matter of the brain. Similarly, hyperphosphorylation of tau, a protein responsible for stabilizing microtubules mainly in neurons of the CNS, at its serine, threonine, and tyrosine residues results in the aggregation of phospho-tau to form intraneuronal neurofibrillary tangles. Studies have also shown that aggregation of α-synuclein, a protein responsible for regulating synaptic vesicle trafficking and inducing synaptic plasticity, contributes to the formation of amyloid β plaques and neurofibrillary tau tangles^2-3^.

However, the biomechanisms involved in the contribution of certain pathologies to Alzheimer’s disease have not been fully determined. In addition, the expression of different genes associated with the disease is an area of interest. This is because the roles various genes play in disease progression have yet to be fully determined. The expression of these disease biomarkers can be compared across brain regions as well as between different subject groups, to determine any notable differences between the gene expression profiles of different brain regions. This information will serve important to the understanding of the disease on a genetic basis, thereby allowing for the development of more targeted pharmacological treatments.

To analyze differences in the expression of genes, we used a curated dataset from the Aging, Dementia, and TBI study conducted by the Allen Brain Institute for Brain Science^4^. These datasets provided post-mortem RNA-sequencing data on the expression of numerous genes in different subjects, as well as subject information such as the presence of the APOE4 allele and Alzheimer’s status. We divided the subject data based on several criteria, such as the brain region in which the gene was expressed, and conducted statistical tests to determine differences in gene expression across different brain regions and sexes.

We found that amyloid β and phospho-tau levels were significantly higher in Alzheimer’s patients who carried the APOE4 allele. Additionally, our results indicate that differences in BDNF expression, amyloid β plaque deposition, and level of oxidative damage between the HIP, FWM, PCx, and TCx brain regions are statistically significant. We also discovered that unlike amyloidogenesis, phospho-tau accumulation has a strong association with sex, with significantly higher Tau-2 immunoreactivity levels in females. These results provide insight on the biochemical factors that play a role in the etiology and development of Alzheimer’s disease.

## Materials and Methods

### Data

RNA sequencing data used in this study was obtained from the Aging, Dementia, and TBI study conducted by the Allen Brain Institute for Brain Science (http://aging.brain-map.org/)^4^. This dataset, derived from the Adult Changes in Thought (ACT) cohort, contains transcriptomic and neuropathologic data collected from a group of 107 subjects. Gene expression data was collected from tissue samples of the subjects’ postmortem brains across four brain regions: the hippocampus, frontal white matter, parietal cortex, and temporal cortex. Luminex assays were used to determine concentrations of protein biomarkers such as amyloid β peptides, forms of tau and phospho-tau, α-synuclein, neurotrophic factors, and inflammatory mediators. In addition, gas chromatography mass spectrometry (GC/MS) analysis of isoprostanes was used to mark and quantify oxidative damage caused by free radicals^4^. The Seaborn, pandas, SciPy, NumPy, and statsmodels Python packages were used for data manipulation and statistical analysis.

### Statistical Analysis

#### T-test

The subjects were separated into several groups: those with dementia caused by Alzheimer’s Disease, vascular dementia, other kinds of dementia, and those without dementia. Alzheimer’s patients were then also divided based on categories such as the presence of the APOE4 allele and sex. Concentrations of biomarkers were compared across these groups using t-tests to determine any significant differences.

### ANOVA

In this study, two-way analysis of variance (ANOVA) tests were used to determine how the APOE4 allele, the brain region from which the tissue was collected, and the patient’s sex have an effect on the concentration of various biomarkers. It also determined any interaction between these three factors.

## Results

Brain-derived neurotrophic factor, or BDNF, plays an important role in neuroplasticity, neurogenesis, learning, and long-term memory^9^. A decrease in BDNF levels has been associated with several neurodegenerative disorders including Alzheimer’s disease^8^. APOE4 is an allele of the apolipoprotein E gene. Studies have shown that carriers of this allele are more likely to develop late-onset Alzheimer’s disease^1^. To investigate the link between BDNF and the APOE4 allele with Alzheimer’s disease pathology, the effect of various factors on the concentration of biomarkers in the brains with Alzheimer’s disease was compared across the four brain regions. A violin plot shows that APOE4 allele influences BDNF (brain-derived neurotrophic factor) concentration, measured as pg of BDNF per mg of total protein, across the four different brain regions (Figure 1).

**Figure 1.**
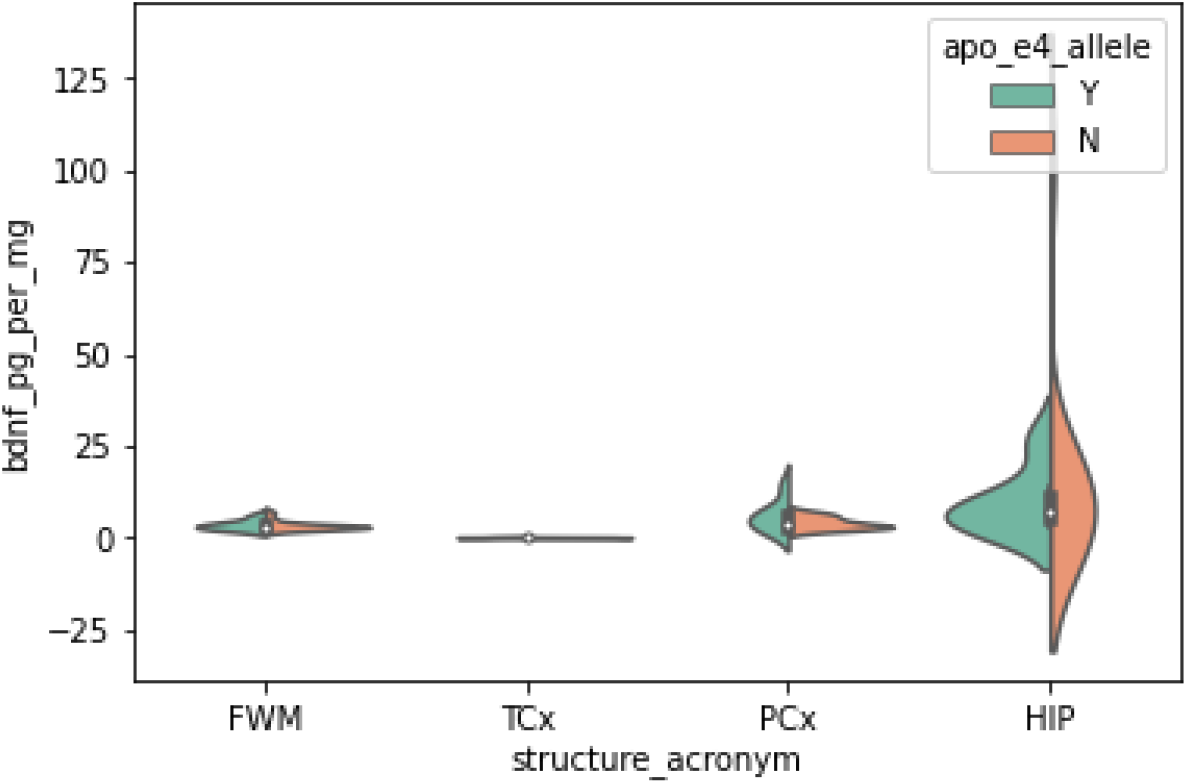
BDNF levels across brain regions. BDNF levels are shown as pg per mg across the four brain regions of interest: FWM, TCx, PCx, and HIP. BDNF levels are further segmented by whether APOE4 allele was present in the patient sample.

A two-way ANOVA (Allele x Structure) showed a non-significant interaction between APOE4 allele presence and brain region (P > 0.5) (Table 1). No statistically significant relationship was found between the presence of APOE4 allele and BDNF concentration (P > 0.5), and follow-up post-hoc analyses via t-test validated this and indicated a non-significant difference between patient groups in hippocampal (P = 0.61), frontal white matter (P = 0.83), and parietal cortex (P = 0.20) BDNF levels. However, there was a statistically significant main effect of brain region (F = 5.19, P < 0.005), indicating a strong correlation between the brain region sample and BDNF concentration, such that BDNF concentration levels appeared to be larger in the hippocampus compared to other regions. We conducted t-tests across brain regions to further investigate this correlation. There was a statistically significant difference in BDNF levels between tissues from the hippocampus and frontal white matter (t = 2.39, P = 0.02), hippocampus and parietal cortex (t = 2.17, P = 0.03), and frontal white matter and parietal cortex (t = -2.10, P = 0.04). Analysis of temporal cortex BDNF expression was not conducted as there was not enough data available. Results suggest that BDNF is differentially expressed across brain regions, suggesting that neuronal growth processes may be brain region-dependent.

**Table 1.** Statistical results comparing BDNF levels across the four brain regions of interest in the presence or absence of the APOE4 allele.

|  | sum_sq | df | F | PR(>F) |
| --- | --- | --- | --- | --- |
| **C(apo_e4_allele)** | 8.933095 | 1.0 | 0.075368 | 0.784411 |
| **C(structure_acronym)** | 1845.865548 | 3.0 | 5.191165 | 0.002556 |
| **C(apo_e4_allele):C(structure_acronym)** | 146.726424 | 3.0 | 0.412642 | 0.744383 |
| **Residual** | 9126.509235 | 77.0 | NaN | NaN |

Another biomarker of interest was amyloid β, as amyloid β plaques are notable histopathologies found in Alzheimer’s tissues^17^. Amyloid β levels were measured as the percentage of fresh frozen tissue area covered by Aβ40 and Aβ42 immunoreactivity. The effect of various factors on amyloid β accumulation in Alzheimer’s brains was compared across the four brain regions. We first observed a differential pattern of Aβ peptide concentration across the four different brain regions depending on whether APOE4 allele was present or not in the subject sample (Figure 2).

**Figure 2.**
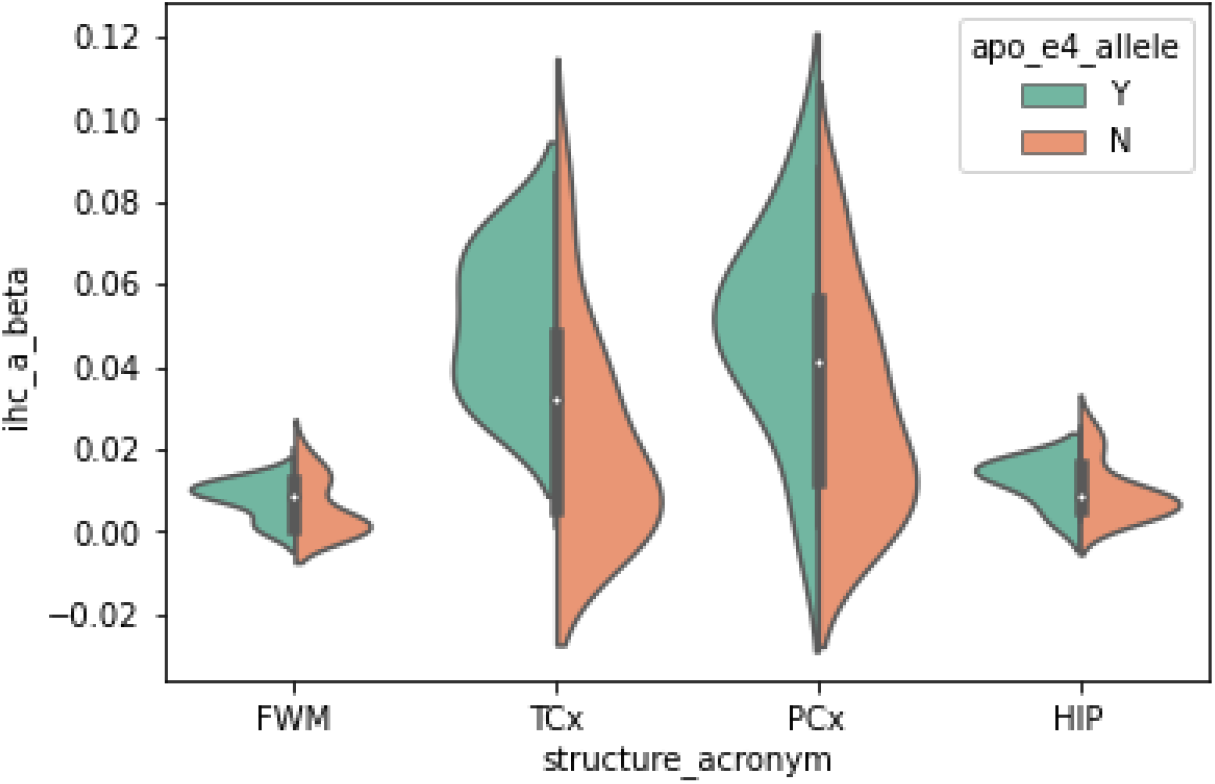
Aβ levels across brain regions. Aβ levels are shown as the percentage of area covered by Aβ40 and Aβ42 immunoreactivity across the four brain regions of interest: FWM, TCx, PCx, and HIP. Aβ levels are further segmented by whether APOE4 allele was present in the patient sample.

A two-way ANOVA (Allele x Structure) appeared to show a trend for a statistically significant interaction between APOE4 allele presence and brain region on amyloid β levels (F = 2.62, P = 0.0558) (Table 2). There was a statistically significant relationship between the presence of APOE4 allele and amyloid β concentration (F = 12.07, P < 0.001). Post-hoc t-test analyses exploring this relationship only showed a statistically significant difference between patient groups in temporal cortex amyloid β levels (P = 0.01). There was a non-significant difference between hippocampal (P = 0.44), frontal white matter (P = 0.76), and parietal cortex (P = 0.15) groups in Aβ levels. There was also a statistically significant main effect of brain region (F = 15.6, P < 0.001), indicating a strong correlation between the brain region sample and degree of Aβ accumulation. Results suggest that amyloid β levels vary significantly across different brain regions, and sometimes based on the presence of the APOE4 allele.

**Table 2.**
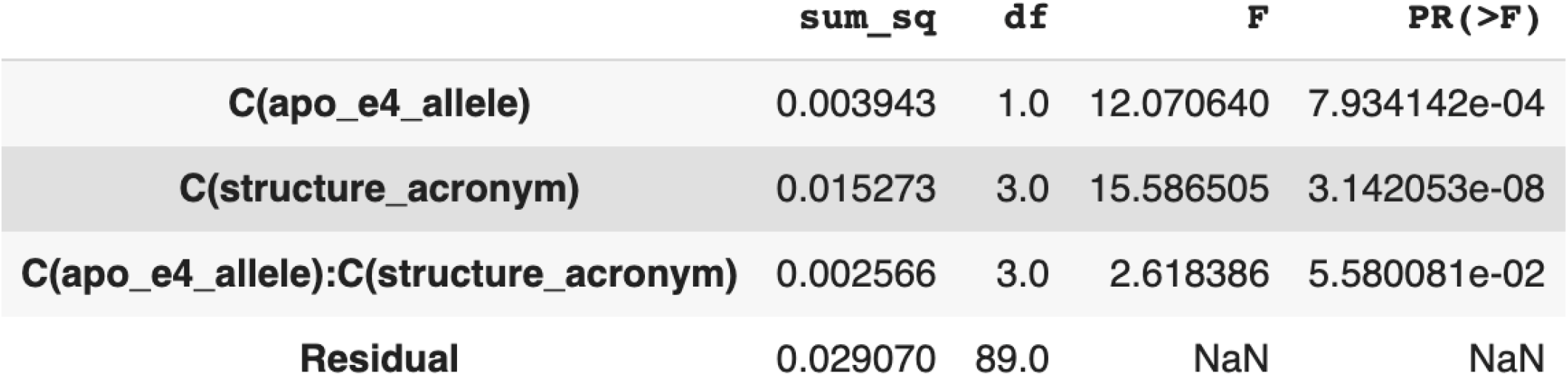
Statistical results comparing Aβ levels across the four brain regions of interest in the presence or absence of the APOE4 allele.

Several studies have shown that the incidence of Alzheimer’s disease is significantly higher in females when compared to their male counterparts^20^. We analyzed the effect of the Alzheimer’s patient’s sex on amyloid β concentration to determine the presence of a biochemical basis behind this finding. We observed that the presence of the APOE4 allele on Aβ peptide concentration across male and female patient groups appeared to differ (Figure 3).

**Figure 3.**
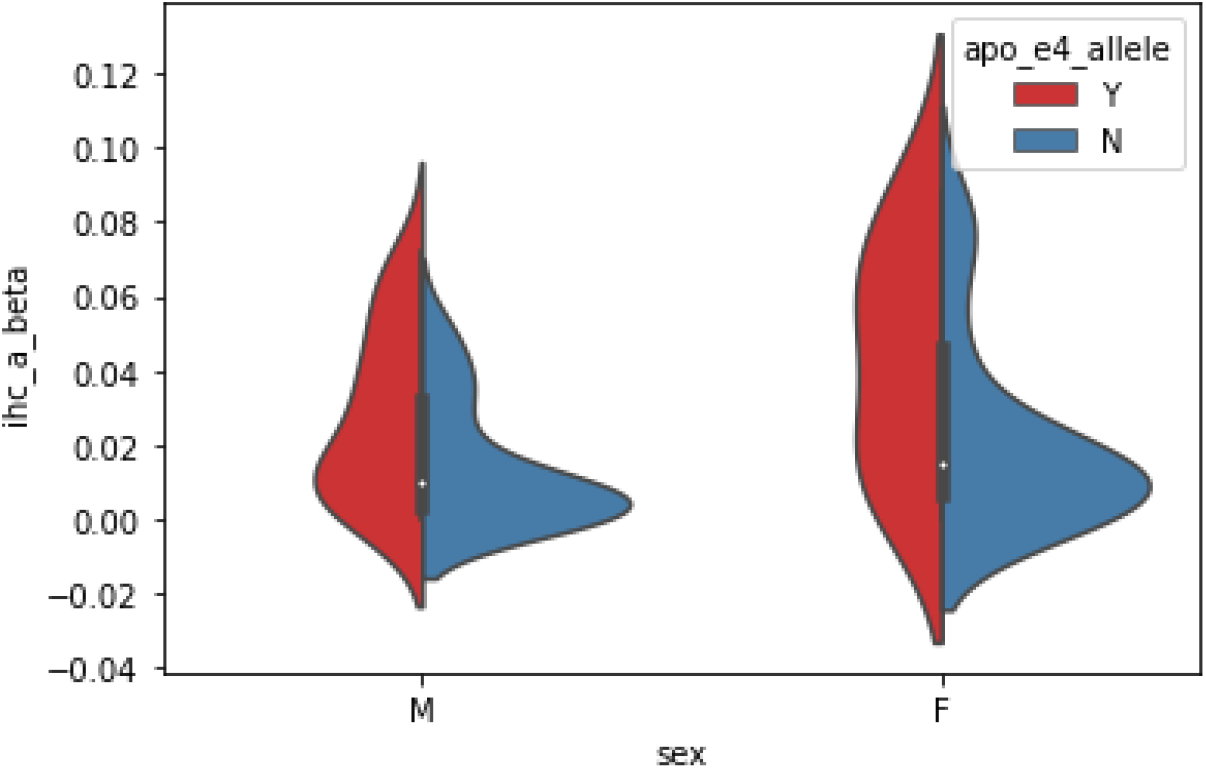
Aβ levels in males vs. females. Aβ levels are shown as the percentage of area covered by Aβ40 and Aβ42 immunoreactivity in males and females. Aβ levels are further segmented by whether APOE4 allele was present in the patient sample.

**Figure 4.**
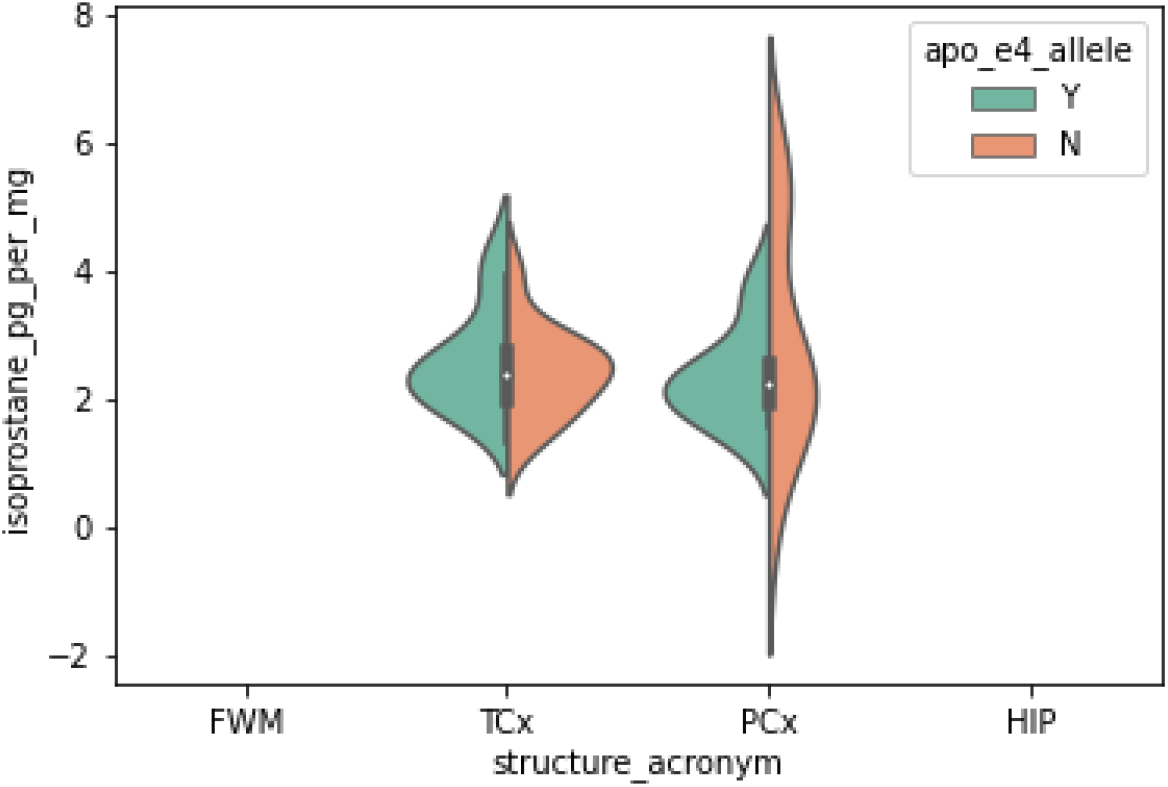
Isoprostane levels across brain regions. Isoprostane levels are shown as pg per mg only across two brain regions: TCx and PCx due to a lack of isoprostane data in the FWM and HIP. BDNF levels are further segmented by whether APOE4 allele was present in the patient sample.

**Figure 5.**
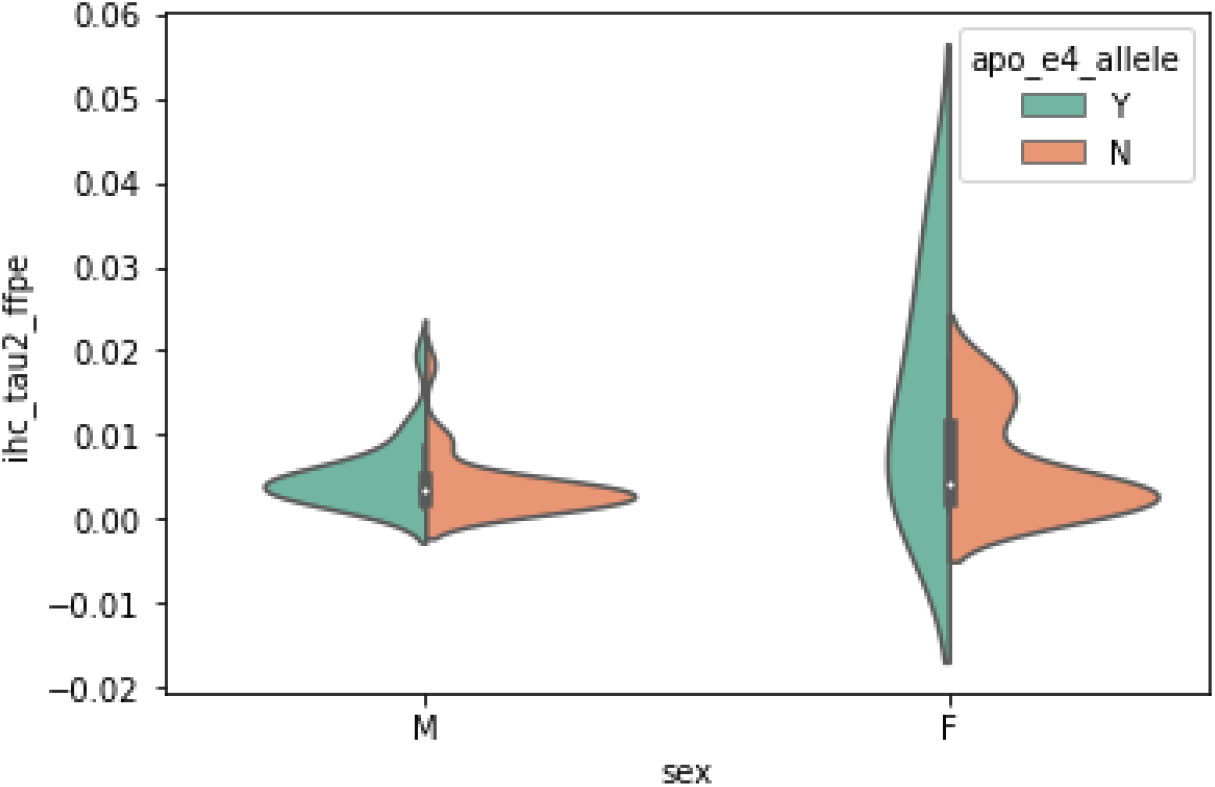
Tau-2 levels in males vs. females. Tau-2 levels are shown as the percentage of area covered by Tau-2 immunoreactivity in males and females. Tau-2 levels are further segmented by whether APOE4 allele was present in the patient sample.

A two-way ANOVA (Allele x Sex) showed a non-significant interaction between APOE4 allele presence and sex on amyloid β levels (P = 0.40) (Table 3). However, there was a statistically significant main effect of APOE4 allele presence on amyloid β concentration (F = 10.81, P = 0.001). Follow-up t-tests revealed a statistically significant difference in Aβ levels between APOE4+ and APOE4-groups divided by sex: male (P < 0.01) and female (P < 0.05). The ANOVA also hinted a statistically significant main effect of sex on amyloid β concentrations (F = 3.92, P = 0.05), indicating the possibility of a correlation between the patient’s sex and Aβ accumulation. However, a post-hoc t-test indicated the lack of a statistically significant difference in Aβ levels between male and female APOE4+ groups (P = 0.12), implying the lack of a strong association between sex and levels of Aβ plaque deposition in Alzheimer’s patients.

**Table 3.** Statistical results comparing Aβ levels across sexes in the presence or absence of the APOE4 allele.

|  | sum_sq | df | F | PR(>F) |
| --- | --- | --- | --- | --- |
| **C(apo_e4_allele)** | 0.005198 | 1.0 | 10.817426 | 0.001421 |
| **C(sex)** | 0.001884 | 1.0 | 3.920585 | 0.050654 |
| **C(apo_e4_allele):C(sex)** | 0.000337 | 1.0 | 0.701430 | 0.404451 |
| **Residual** | 0.044687 | 93.0 | NaN | NaN |

The next biomarkers examined in this study were F2-isoprostanes. F2-isoprostanes are prostaglandin-like compounds produced by the nonenzymatic reaction of free-radicals with arachidonic acid. They are considered to be reliable biomarkers of oxidative stress^6^. An increase in the concentrations of F2-isoprostanes has been associated with Alzheimer’s disease^7^. To investigate the link between isoprostanes and the APOE4 allele with Alzheimer’s disease pathology, the effect of APOE4 allele presence on the concentration of F2-isoprostanes in the brains with Alzheimer’s disease was compared across four different brain regions. We observed that the effect of the APOE4 allele on F2-isoprostane concentration, measured as pg of F2-isoprostane per mg of total protein, across the parietal and temporal cortex, appeared to differ. Analyses were constrained to these two regions due to a lack of data regarding F2-isoprostane concentration in the hippocampus and frontal white matter.

A two-way ANOVA (Allele x Structure) indicated a statistically non-significant interaction between APOE4 allele presence and brain region on F2-isoprostane levels (F = 0.43, P = 0.66) (Table 4). It also showed the lack of a statistically significant relationship between the presence of the APOE4 allele and F2-isoprostane concentration (F = 0.12, P = 0.73). The ANOVA indicates a statistically significant main effect of the brain region (F = 90.7, P << 0.001). However, this figure may be exaggerated as the statistical test may have been influenced by the lack of lipidomic data in the FWM and HIP. Post-hoc analyses using a t-test failed to show a significant difference between F2-isoprostane levels in temporal and parietal cortex groups (P = 0.59). This suggests that the degree of oxidative damage done in tissues due to Alzheimer’s does not vary significantly based on brain region.

**Table 4.**
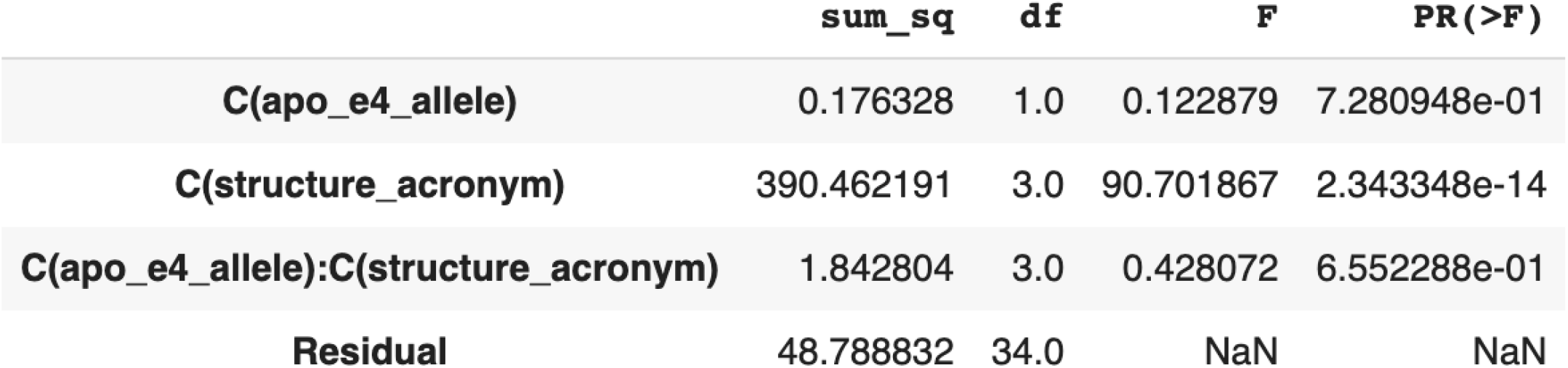
Statistical results comparing isoprostane levels across the two brain regions in the presence or absence of the APOE4 allele.

|  | sum_sq | df | F | PR(>F) |
| --- | --- | --- | --- | --- |
| **C(apo_e4_allele)** | 0.176328 | 1.0 | 0.122879 | 7.280948e-01 |
| **C(structure_acronym)** | 390.462191 | 3.0 | 90.701867 | 2.343348e-14 |
| **C(apo_e4_allele):C(structure_acronym)** | 1.842804 | 3.0 | 0.428072 | 6.552288e-01 |
| **Residual** | 48.788832 | 34.0 | NaN | NaN |

**Table 5.**
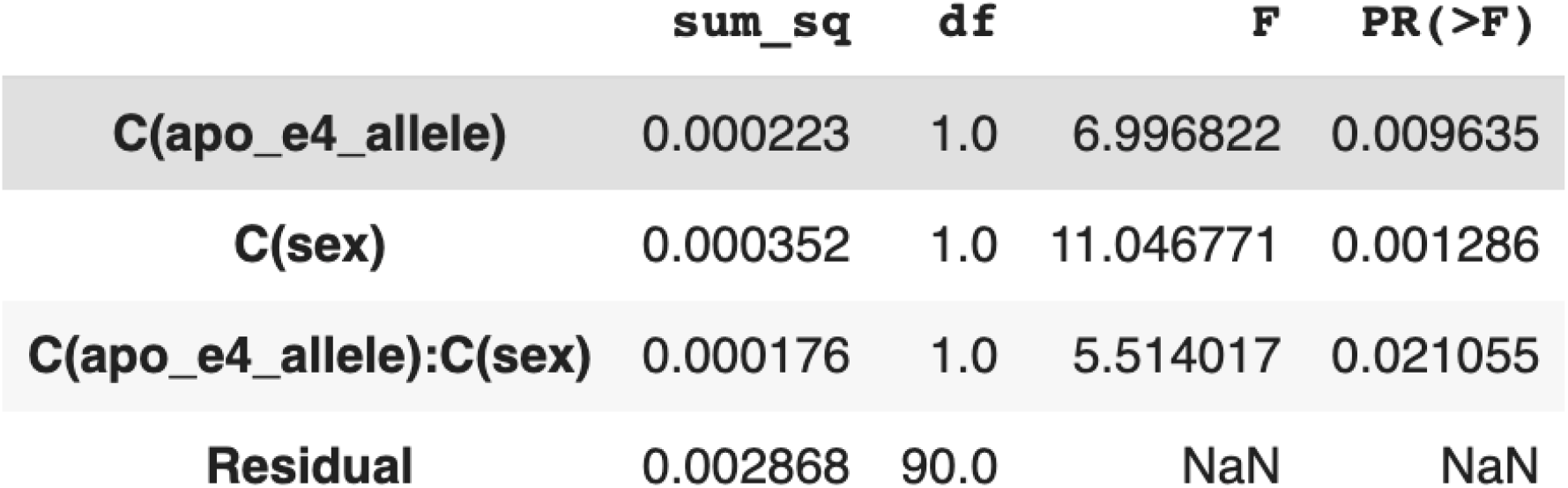
Statistical results comparing Tau-2 levels across sexes in the presence or absence of the APOE4 allele.

|  | sum_sq | df | F | PR(>F) |
| --- | --- | --- | --- | --- |
| **C(apo_e4_allele)** | 0.000223 | 1.0 | 6.996822 | 0.009635 |
| **C(sex)** | 0.000352 | 1.0 | 11.046771 | 0.001286 |
| **C(apo_e4_allele):C(sex)** | 0.000176 | 1.0 | 5.514017 | 0.021055 |
| **Residual** | 0.002868 | 90.0 | NaN | NaN |

The final biomarker analyzed in this study was Tau-2 immunoreactivity, measured as the percentage of tissue area covered. Tau-2 is a BODIPY-based fluorescent probe synthetically designed to identify hyperphosphorylated tau proteins^35^. In this study, the Tau-2 biomarker was used to identify mature neurofibrillary tau tangles and dystrophic neurites. Studies have shown that women have more neurofibrillary tangles and AD-associated pathology^24^. We analyzed the effect of the Alzheimer’s patient’s sex on Tau-2 concentration to also determine a link with APOE4 carrier status. We observed that the presence of the APOE4 allele appeared to result in differential Tau-2 concentration levels across male and female patient groups.

A two-way ANOVA (Allele x Sex) showed a statistically significant interaction between APOE4 allele presence and sex on levels of Tau-2 immunoreactivity (F = 5.51, P = 0.02). There was a statistically significant relationship between the presence of APOE4 allele and Tau-2 concentration (F = 7.00, P < 0.01). Post-hoc analyses via t-tests only showed a statistically significant difference in Tau-2 levels between female APOE4+/-patient groups (P = 0.02). There was no statistically significant difference between male APOE4+/-groups (P = 0.16). There was also a statistically significant main effect of sex (F = 11.05, P = 0.001), indicating a strong correlation between the patient’s sex and degree of Tau-2 accumulation. Results suggest that Tau-2 immunoreactivity varies in concentration significantly based on sex, and in females only, on the presence of the APOE4 allele. This implies that sex likely influences the degree of aggregation of hyperphosphorylated tau proteins and the formation of neurofibrillary tangles.

## Discussion

The etiologies of Alzheimer’s disease are not fully understood, especially on a genetic level. This makes early diagnosis and treatment of the disease more difficult. However, gene expression and proteomic analysis can reveal biochemical changes in certain diseases, allowing for the identification of specific molecular and histological biomarkers as well as possible therapeutic targets. In this study, we analyzed transcriptomic and proteomic data of Alzheimer’s patients across multiple brain regions implicated in Alzheimer’s pathology and cognitive symptom manifestation. Indeed, although the analyzed brain regions vary in structure and function, transcriptional changes in these regions have been previously connected to the pathogenesis of Alzheimer’s disease. In addition, these four brain regions cover a large area of the brain, allowing for AD-associated changes to be differentiated throughout the brain and capture of region-specific changes in AD-associated biomarkers.

Analyses determined that there was a significant difference between groups of different brain regions in BDNF expression levels, consistent with prior reports^8^. BDNF is a vital neurotrophin responsible for regulating neural plasticity, growth, differentiation, and synaptic strength^9^. These neurological processes are essential for higher-level thinking, cognition, and most notably, memory^10,11^. Learning and memory are primary functions of the hippocampus^12^, a plastic structure, which may explain the increased BDNF expression levels observed in the region. BDNF preserves microstructural integrity in white matter^14^, and also serves an important role in the cerebral cortex, primarily responsible for cognitive processes such as learning and decision-making^13^. Analysis showed that there was no significant relationship between presence of the APOE4 allele and BDNF expression in Alzheimer’s patients. However, a previous study reported that APOE4 allele carriers showed lower levels of BDNF^15^. This is logical as APOE4 carriers have shown to be more susceptible to developing Alzheimer’s disease^1^, and reduced levels of neurotrophins, such as BDNF, have been strongly associated with AD symptoms and pathogenesis^8^. The weak correlation found between APOE4 allele presence and BDNF levels in our study is likely due to the relatively small sample size of the original Aging, Dementia, and TBI study^4^.

Analysis of the effect of APOE4 allele on amyloid β levels showed a strong correlation between the two variables. Those carrying the APOE4 allele often had greater amounts of amyloid β in their brain tissue—a phenomenon that was most apparent in the temporal cortex. Prior work has shown that APOE4 presence is linked more with temporal atrophy than with frontal atrophy^16^. Similarly, amyloid β accumulation has been linked to brain atrophy, especially in the temporoparietal regions^17^. Therefore, the greater increase in amyloid β concentration in the temporal cortex, in comparison to other brain regions, may be due to the presence of the APOE4 allele. Our study also showed differential amyloid β levels across the brain regions of interest, which may have been due to the unique spatial affinities of amyloid β to specific brain tissue compositions, such as the abundance of white matter or cell bodies.

Next, analysis of the effect of sex on amyloid β concentration indicated the possibility of a significant relationship (P = 0.05). Female Alzheimer’s patients seemed to often have greater amounts of amyloid β levels when compared to male patients. A study reported that an increase in mitochondrial generation of reactive oxygen species has been associated with an increase in amyloid β concentration^20^. In young females, upregulation of antioxidant genes by estrogens and phytoestrogens has been shown to effectively subdue the free radicals, thereby limiting oxidative stress^20,21^. However, with aging and menopause, females around their 50s start to experience a decrease in estrogen levels^22^. This would explain a decrease in protection against Aβ-induced oxidative damage, accounting for greater amyloid β concentrations in females. This directly links to the fact that Alzheimer’s disease is more prevalent among women than men^22^.

Our study also determined statistically significant differences in F2-isoprostane concentrations across different brain regions. A previous gene expression study suggested that differences in degree of oxidative damage caused by reactive oxygen species account for differences in intensity of neurodegeneration in different brain regions^23^. As F2-isoprostanes serve as reliable biomarkers of such oxidative damage^6^, it can be concluded that brain regions with greater concentrations of F2-isoprostanes are affected more by Alzheimer’s disease. This idea warrants further research into the question as to which brain regions are more susceptible to AD-induced oxidative damage and why.

Analysis of Tau-2 immunoreactivity levels revealed a statistically significant difference between APOE4+/-patient groups (P < 0.01). Previous studies have also shown that APOE4 increases accumulation of tau in the brain^25,26,27^. One study proposed that the N-terminal domain generated by the proteolysis of the APOE4 protein promotes the hyperphosphorylation and accumulation of tau^28^. It has also been shown that APOE4 leads to the upregulation of C/EBPβ, which serves as a transcription factor for δ-secretase—the enzyme that cleaves APP, tau, α-synuclein, and a multitude of other proteins^29,30,31^. This may explain how the APOE4 allele is correlated with increased tau accumulation. Our study also showed a significant difference in Tau-2 levels between males and females (P = 0.001). Prior studies have shown that testosterone may have a neuroprotective effect against phospho-tau in Alzheimer’s patients, especially APOE4 carriers^32,32^. Some have also shown that estrogen interacts with tau to inhibit hyperphosphorylation and aggregation^34^.

Therefore, lower testosterone levels in females and decrease in estrogen levels with age may explain the increased phospho-tau levels. However, further research into how the biochemical pathways associated with tau phosphorylation differ in males and females is necessary to elucidate the relationship between sex and neurofibrillary tangle formation.

Post-mortem tissue studies allow us to increase our understanding of how different genes impact the presentation of Alzheimer’s pathology. Through a data-science approach, we analyzed how different biomarkers were differentially expressed given the presence of APOE4 allele and other demographic factors. These findings provide novel avenues for further studies examining biological processes linked to Alzheimer’s disease dementia.

## Data Availability

All data produced are available online at http://aging.brain-map.org/download/index.

## References

1. Kumar, A., Sidhu, J., Goyal, A., & Tsao, J. W. (2021). Alzheimer Disease. In StatPearls. StatPearls Publishing.

2. Vacchi, E., Kaelin-Lang, A., & Melli, G. (2020). Tau and Alpha Synuclein Synergistic Effect in Neurodegenerative Diseases: When the Periphery Is the Core. International journal of molecular sciences, 21(14), 5030. https://doi.org/10.3390/ijms21145030

3. Crews, L., Tsigelny, I., Hashimoto, M., & Masliah, E. (2009). Role of synucleins in Alzheimer’s disease. Neurotoxicity research, 16(3), 306–317. https://doi.org/10.1007/s12640-009-9073-6

4. Allen Institute for Brain Science: Aging, Dementia and TBI Study

5. Filon, J. R., Intorcia, A. J., Sue, L. I., Vazquez Arreola, E., Wilson, J., Davis, K. J., Sabbagh, M. N., Belden, C. M., Caselli, R. J., Adler, C. H., Woodruff, B. K., Rapscak, S. Z., Ahern, G. L., Burke, A. D., Jacobson, S., Shill, H. A., Driver-Dunckley, E., Chen, K., Reiman, E. M., … Serrano, G. E. (2016). Gender Differences in Alzheimer Disease: Brain Atrophy, Histopathology Burden, and Cognition. Journal of Neuropathology & Experimental Neurology, 75(8), 748–754. https://doi.org/10.1093/jnen/nlw047

6. Montuschi, P., Barnes, P. J., & Roberts, L. J., 2nd (2004). Isoprostanes: markers and mediators of oxidative stress. FASEB journal : official publication of the Federation of American Societies for Experimental Biology, 18(15), 1791–1800. https://doi.org/10.1096/fj.04-2330rev

7. Praticò, D., MY Lee, V., Trojanowski, J. Q., Rokach, J., & Fitzgerald, G. A. (1998). Increased F2-isoprostanes in Alzheimer’s disease: evidence for enhanced lipid peroxidation in vivo. FASEB journal : official publication of the Federation of American Societies for Experimental Biology, 12(15), 1777–1783. https://doi.org/10.1096/fasebj.12.15.1777

8. Devlin, P., Cao, X., & Stanfill, A. G. (2021). Genotype-expression interactions for BDNF across human brain regions. BMC Genomics, 22(1). https://doi.org/10.1186/s12864-021-07525-1

9. Huang, E. J., & Reichardt, L. F. (2001). Neurotrophins: roles in neuronal development and function. Annual review of neuroscience, 24, 677–736. https://doi.org/10.1146/annurev.neuro.24.1.677

10. Yamada, K., & Nabeshima, T. (2003). Brain-Derived Neurotrophic Factor/TrkB Signaling in Memory Processes. Journal of Pharmacological Sciences, 91(4), 267–270. https://doi.org/10.1254/jphs.91.267

11. Cavaleiro, C., Martins, J., Gonçalves, J., & Castelo-Branco, M. (2020). Memory and Cognition-Related Neuroplasticity Enhancement by Transcranial Direct Current Stimulation in Rodents: A Systematic Review. Neural plasticity, 2020, 4795267. https://doi.org/10.1155/2020/4795267

12. Anand, K. S., & Dhikav, V. (2012). Hippocampus in health and disease: An overview. Annals of Indian Academy of Neurology, 15(4), 239–246. https://doi.org/10.4103/0972-2327.104323

13. Jawabri, K. H., & Sharma, S. (2021). Physiology, Cerebral Cortex Functions. In StatPearls. StatPearls Publishing.

14. Maillard, P., Satizabal, C.L., Beiser, A.S., Himali, J.J., Chen, T., Vasan, R.S., Seshadri, S., & DeCarli, C. (2016). Effects of Brain Derived Neurotrophic Factor on White Matter Integrity in Middle-Aged Adults: A Voxel-Based Diffusion Tensor Imaging Study (S39.006). Neurology, 86.

15. Alvarez, A., Aleixandre, M., Linares, C., Masliah, E., & Moessler, H. (2014). Apathy and APOE4 are associated with reduced BDNF levels in Alzheimer’s disease. Journal of Alzheimer’s disease : JAD, 42(4), 1347–1355. https://doi.org/10.3233/JAD-140849

16. Geroldi, C., Pihlajamäki, M., Laakso, M. P., DeCarli, C., Beltramello, A., Bianchetti, A., Soininen, H., Trabucchi, M., & Frisoni, G. B. (1999). APOE-epsilon4 is associated with less frontal and more medial temporal lobe atrophy in AD. Neurology, 53(8), 1825–1832. https://doi.org/10.1212/wnl.53.8.1825

17. Palmqvist, S., Schöll, M., Strandberg, O., Mattsson, N., Stomrud, E., Zetterberg, H., Blennow, K., Landau, S., Jagust, W., & Hansson, O. (2017). Earliest accumulation of β-amyloid occurs within the default-mode network and concurrently affects brain connectivity. Nature Communications, 8(1). https://doi.org/10.1038/s41467-017-01150-x

18. Pereira, J. B., Ossenkoppele, R., Palmqvist, S., Strandberg, T. O., Smith, R., Westman, E., & Hansson, O. (2019). Amyloid and tau accumulate across distinct spatial networks and are differentially associated with brain connectivity. eLife, 8, e50830. https://doi.org/10.7554/eLife.50830

19. Braak, H., & Braak, E. (1991). Neuropathological stageing of Alzheimer-related changes. Acta neuropathologica, 82(4), 239–259. https://doi.org/10.1007/BF00308809

20. Viña, J., & Lloret, A. (2010). Why women have more Alzheimer’s disease than men: gender and mitochondrial toxicity of amyloid-beta peptide. Journal of Alzheimer’s disease : JAD, 20 Suppl 2, S527–S533. https://doi.org/10.3233/JAD-2010-100501

21. Horstman, A. M., Dillon, E. L., Urban, R. J., & Sheffield-Moore, M. (2012). The Role of Androgens and Estrogens on Healthy Aging and Longevity. The Journals of Gerontology Series A: Biological Sciences and Medical Sciences, 67(11), 1140–1152. https://doi.org/10.1093/gerona/gls068

22. Andrew, M. K., & Tierney, M. C. (2018). The puzzle of sex, gender and Alzheimer’s disease: Why are women more often affected than men?. Women’s Health, 14, 1745506518817995. https://doi.org/10.1177/1745506518817995

23. Aksenov, M. Y., Tucker, H. M., Nair, P., Aksenova, M. V., Butterfield, D. A., Estus, S., & Markesbery, W. R. (1998). The expression of key oxidative stress-handling genes in different brain regions in Alzheimer’s disease. Journal of molecular neuroscience : MN, 11(2), 151–164. https://doi.org/10.1385/JMN:11:2:151

24. Barnes, L. L., Wilson, R. S., Bienias, J. L., Schneider, J. A., Evans, D. A., & Bennett, D. A. (2005). Sex differences in the clinical manifestations of Alzheimer disease pathology. Archives of general psychiatry, 62(6), 685–691. https://doi.org/10.1001/archpsyc.62.6.685

25. Baek, M. S., Cho, H., Lee, H. S., Lee, J. H., Ryu, Y. H., & Lyoo, C. H. (2020). Effect of APOE ε4 genotype on amyloid-β and tau accumulation in Alzheimer’s disease. Alzheimer’s Research & Therapy, 12(1). https://doi.org/10.1186/s13195-020-00710-6

26. Shi, Y., Yamada, K., Liddelow, S. A., Smith, S. T., Zhao, L., Luo, W., Tsai, R. M., Spina, S., Grinberg, L. T., Rojas, J. C., Gallardo, G., Wang, K., Roh, J., Robinson, G., Finn, M. B., Jiang, H., Sullivan, P. M., Baufeld, C., Wood, M. W., Sutphen, C., … Holtzman, D. M. (2017). ApoE4 markedly exacerbates tau-mediated neurodegeneration in a mouse model of tauopathy. Nature, 549(7673), 523–527. https://doi.org/10.1038/nature24016

27. Zhou, M., Huang, T., Collins, N., Zhang, J., Shen, H., Dai, X., Xiao, N., Wu, X., Wei, Z., York, J., Lin, L., Zhu, Y., LaDu, M. J., & Chen, X. (2016). APOE4 Induces Site-Specific Tau Phosphorylation Through Calpain-CDK5 Signaling Pathway in EFAD-Tg Mice. Current Alzheimer research, 13(9), 1048–1055. https://doi.org/10.2174/1567205013666160415154550

28. Rohn T. T. (2013). Proteolytic cleavage of apolipoprotein E4 as the keystone for the heightened risk associated with Alzheimer’s disease. International journal of molecular sciences, 14(7), 14908–14922. https://doi.org/10.3390/ijms140714908

29. Wang, Z. H., Xia, Y., Liu, P., Liu, X., Edgington-Mitchell, L., Lei, K., Yu, S. P., Wang, X. C., & Ye, K. (2021). ApoE4 activates C/EBPβ/δ-secretase with 27-hydroxycholesterol, driving the pathogenesis of Alzheimer’s disease. Progress in neurobiology, 202, 102032. https://doi.org/10.1016/j.pneurobio.2021.102032

30. Wang, Z. H., Xiang, J., Liu, X., Yu, S. P., Manfredsson, F. P., Sandoval, I. M., Wu, S., Wang, J. Z., & Ye, K. (2019). Deficiency in BDNF/TrkB Neurotrophic Activity Stimulates δ-Secretase by Upregulating C/EBPβ in Alzheimer’s Disease. Cell reports, 28(3), 655–669.e5. https://doi.org/10.1016/j.celrep.2019.06.054

31. Zhang, Z., Tian, Y., & Ye, K. (2020). δ-secretase in neurodegenerative diseases: mechanisms, regulators and therapeutic opportunities. Translational neurodegeneration, 9, 1. https://doi.org/10.1186/s40035-019-0179-3

32. Sundermann, E. E., Panizzon, M. S., Chen, X., Andrews, M., Galasko, D., & Banks, S. J. (2020). Sex differences in Alzheimer’s-related Tau biomarkers and a mediating effect of testosterone. Biology of Sex Differences, 11(1). https://doi.org/10.1186/s13293-020-00310-x

33. Hammond, J., Le, Q., Goodyer, C., Gelfand, M., Trifiro, M., & LeBlanc, A. (2001). Testosterone-mediated neuroprotection through the androgen receptor in human primary neurons. Journal of neurochemistry, 77(5), 1319–1326. https://doi.org/10.1046/j.1471-4159.2001.00345.x

34. Wang, C., Zhang, F., Jiang, S., Siedlak, S. L., Shen, L., Perry, G., Wang, X., Tang, B., & Zhu, X. (2016). Estrogen receptor-α is localized to neurofibrillary tangles in Alzheimer’s disease. Scientific Reports, 6(1). https://doi.org/10.1038/srep20352

35. Verwilst, P., Kim, H. R., Seo, J., Sohn, N. W., Cha, S. Y., Kim, Y., Maeng, S., Shin, J. W., Kwak, J. H., Kang, C., & Kim, J. S. (2017). Rational Design of in Vivo Tau Tangle-Selective Near-Infrared Fluorophores: Expanding the BODIPY Universe. Journal of the American Chemical Society, 139(38), 13393–13403. https://doi.org/10.1021/jacs.7b05878

